# Post-COVID rebound of gonorrhoea in England

**DOI:** 10.1101/2023.08.18.23294107

**Authors:** Holly Fountain, Stephanie Migchelsen, Hannah Charles, Tika Ram, Helen Fifer, Hamish Mohammed, Katy Sinka

## Abstract

With the removal of all COVID-19 restrictions in July 2021, a marked increase in gonorrhoea diagnoses in England was observed. Investigations revealed increases have been widespread, particularly in young people aged 15-to-24 years. Testing numbers have not shown a corresponding increase and have remained below 2019 (pre-pandemic) levels.

**Ethical statement:** No ethical approval was needed for this study as routine surveillance data was used.

**Funding statement:** No funding was needed.

**Conflict of interest:** None

## Background

The COVID-19 pandemic led to widespread social, economic and healthcare service disruption, with substantial disruption to sexual health services (SHSs) in England, including decreased testing. This contributed to a 33.5% (468,260 to 311,480) decrease in new STI diagnoses in 2020 compared to 2019 [1]. From July 2021, all COVID-19 restrictions in England were lifted [2], normal social mixing was permitted and there was a rebound in service provision; a 23.8% (317,022 to 392,453) increase in new STI diagnoses was noted in England between 2021 and 2022 [1]. Of the most commonly diagnosed STIs, the largest proportional increase was for gonorrhoea; compared to 2021, the number of new gonorrhoea diagnoses in 2022 increased by 50.3% (54,961 to 82,592) [1].

The aim of this analysis is to understand trends in tests and diagnoses of gonorrhoea since the lifting of all COVID-19 control measures, and to explore how that differs among different population groups.

## Methods

In England, all STI tests and diagnoses from SHSs are captured by the GUMCAD STI Surveillance System (herein GUMCAD) [3]. Data on gonorrhoea tests and diagnoses between 1 January 2019 and 31 December 2022 were included in this analysis. To prevent double-counting, only one test or diagnosis per individual SHS service user was counted within a 42-day period.

We used these data to examine trends quarterly, and disaggregated by age group (15-24, 25-34, 35-44, 45+ years), gender and sexual orientation (gay, bisexual and other men who have sex with men (GBMSM), heterosexual men, women who have sex with men (WSM), women who have sex with women (WSW)), and local authority (LA) district of residence.

Records with missing data for presented demographic variables were not included in their respective analyses.

Data were analysed using Stata V.16.1 (StataCorp LLC, College Station, Texas, USA).

## Results

Following the lifting of COVID-19 restrictions (Q3 2021) to the end of 2022 (Q4 2022), the number of gonorrhoea tests increased 5.6% (483,717 to 510,792). Gonorrhoea diagnoses, however, increased to a larger extent (63.8%; 13,715 to 22,471) (Figure 1). The total number of diagnoses was the highest on record in 2022, with testing only just returning to 2019 levels.

**Figure 1:**
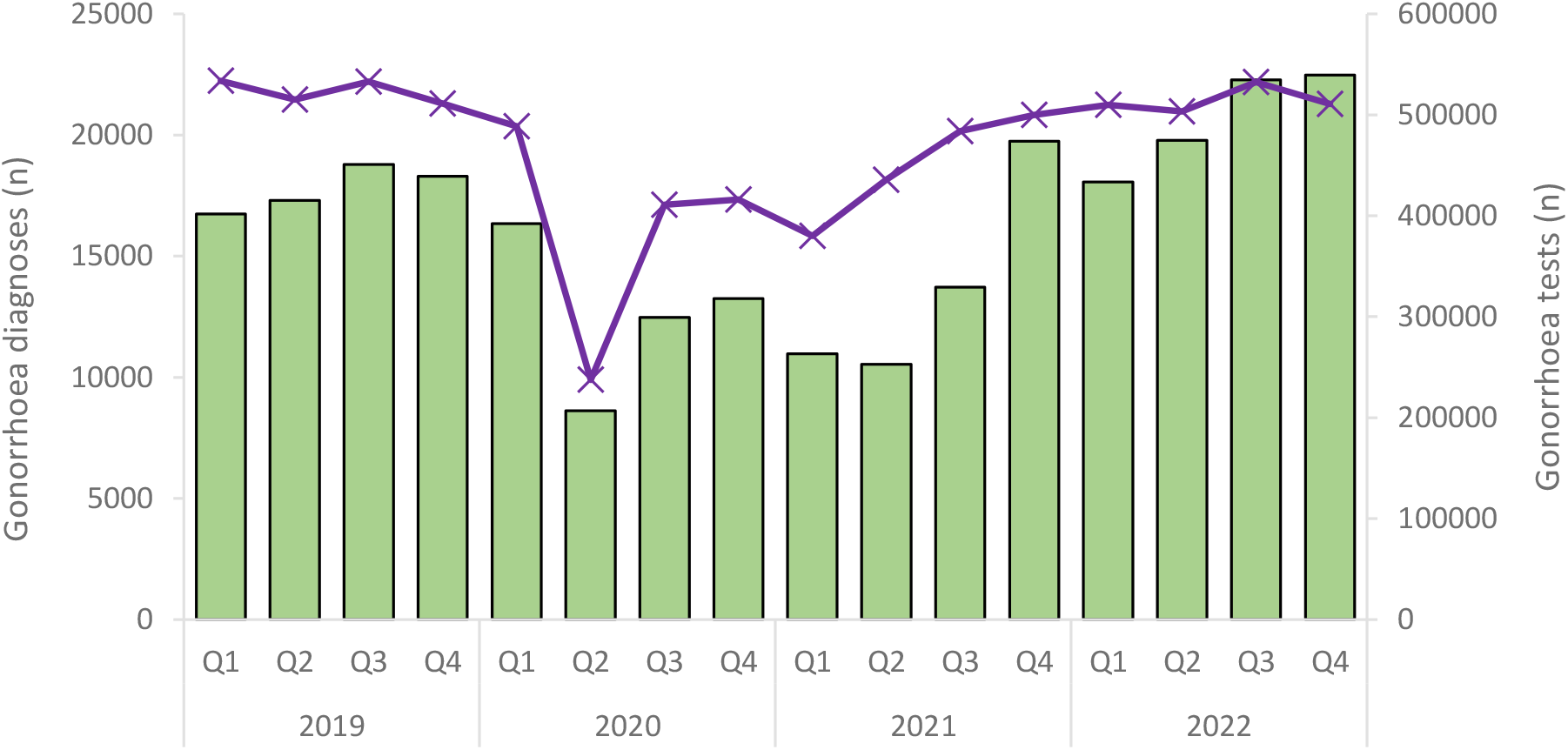
Number of gonorrhoea diagnoses (columns) and gonorrhoea tests (line, secondary axis), England, Quarter 1 2019 to Quarter 4 2022.

The increase in diagnoses began immediately following the lifting of COVID-19 restrictions and was most notable in young people aged 15 to 24 years, where there was a 141.3% increase in diagnoses from Q3 2021 to Q4 2022 (3,747 to 9,041) (Figure 2a), compared to a 34.8% increase for those aged 25 and over. Among young people, 19-to-20-year-olds had the highest increase in diagnoses (229.0%; 930 to 3,060) [data not shown]. In this same period, testing remained steady in young people (1.5% increase). Overall, testing returned to or exceeded 2019 numbers in all age groups except 15-to-24-year-olds (−12.4% Q4 2019 to Q4 2022) (Figure 2b).

**Figure 2:**
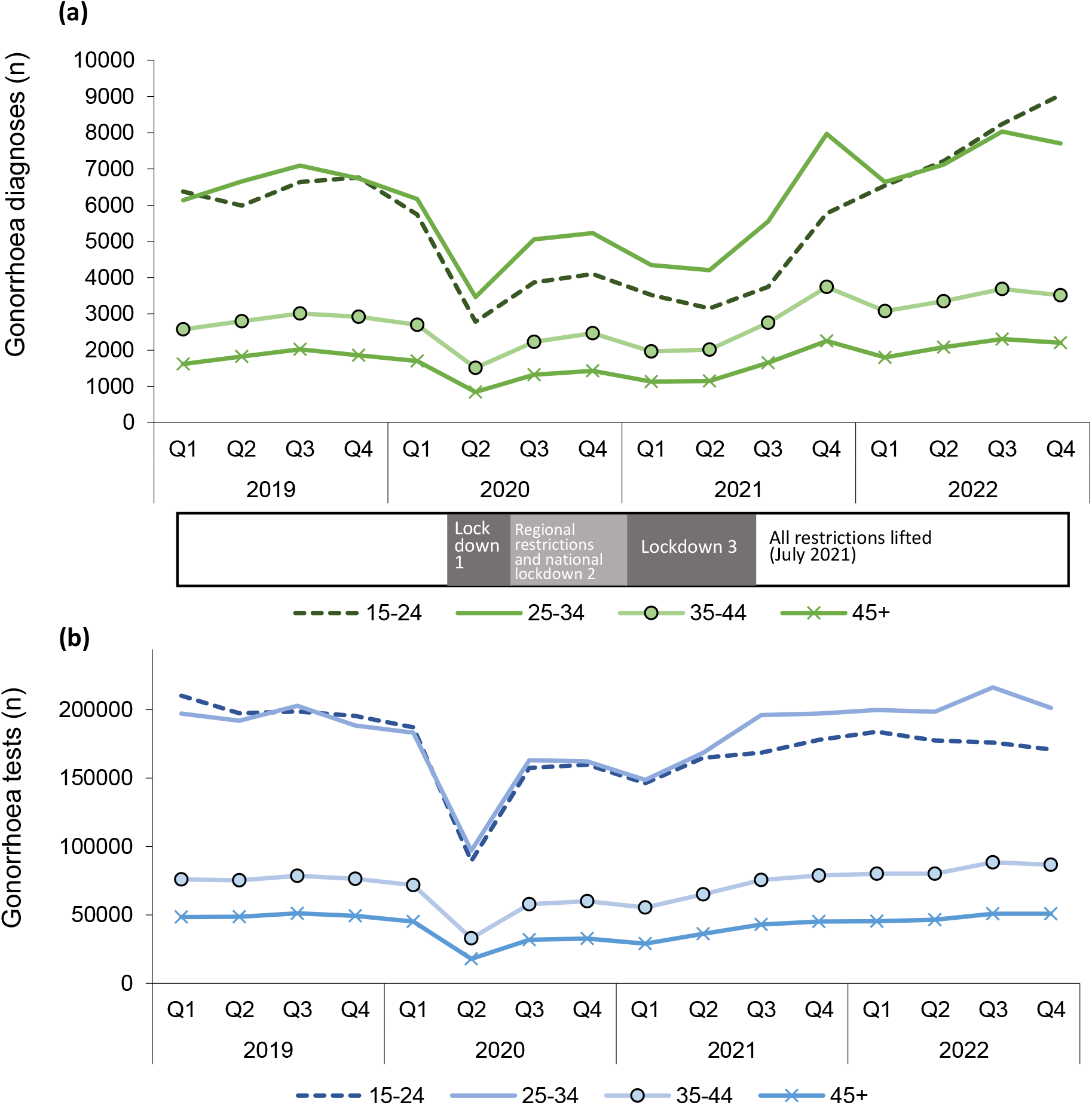
Number of (a) gonorrhoea diagnoses and (b) gonorrhoea tests by age group, England, Quarter 1 2019 to Quarter 4 2022.

Of all gender and sexual orientation groups, GBMSM had the highest amount of gonorrhoea diagnoses. However, proportionally the increase following the lifting of COVID-19 restrictions was largest in WSM (104.7% increase; 2,577 to 5,274), followed by heterosexual men (90.4%; 2,152 to 4,097) and then GBMSM (39.7%; 7,107 to 9,932) (Table 1). Testing increased amongst GBMSM compared to 2019, with numbers up by 21.8% between Q4 2019 and Q4 2022; in this same period however, testing decreased by 20.3% and 8.1% for WSM and heterosexual men respectively [data not shown].

**Table 1:** Number of gonorrhoea diagnoses by gender and sexual orientation, England, Quarter 1 2019 to Quarter 4 2022.

| Year | Quarter | Gonorrhoea diagnoses (n) |  |  |  |  |
| --- | --- | --- | --- | --- | --- | --- |
|  |  | GBMSM | Heterosexual men | WSM | WSW | Not known |
| 2019 | Q1 | 7851 | 3678 | 4280 | 44 | 889 |
|  | Q2 | 8341 | 3692 | 4205 | 64 | 1005 |
|  | Q3 | 9249 | 3832 | 4498 | 55 | 1152 |
|  | Q4 | 8466 | 3909 | 4757 | 47 | 1119 |
| 2020 | Q1 | 8040 | 3356 | 3883 | 60 | 1005 |
|  | Q2 | 4352 | 1681 | 1962 | 39 | 580 |
|  | Q3 | 6194 | 2436 | 2832 | 59 | 956 |
|  | Q4 | 6491 | 2585 | 2964 | 69 | 1134 |
| 2021 | Q1 | 5184 | 2041 | 2392 | 52 | 1305 |
|  | Q2 | 5251 | 1674 | 2076 | 77 | 1456 |
|  | Q3 | 7107 | 2152 | 2577 | 69 | 1810 |
|  | Q4 | 8724 | 2674 | 3665 | 85 | 4590 |
| 2022 | Q1 | 8809 | 2916 | 3809 | 109 | 2419 |
|  | Q2 | 9684 | 3224 | 4182 | 127 | 2560 |
|  | Q3 | 10498 | 3684 | 5032 | 115 | 2953 |
|  | Q4 | 9932 | 4097 | 5274 | 123 | 3045 |

Among 15-to-24-year-olds, the increase in diagnoses from Q3 2021 to Q4 2022 was largest amongst heterosexual people (179.6%; 2,092 to 5,850) compared to GBMSM (63.2%; 1,035 to 1,689). Whilst in those aged 25 and over, diagnoses increased similarly amongst heterosexual people and GBMSM (33.7% and 35.8%, respectively).

Diagnoses of gonorrhoea increased in all regions of England since the removal of COVID-19 restrictions, most notably in the South West (226.0% increase from Q3 2021 to Q4 2022; 407 to 1,327) and the North East (194.0% increase; 285 to 838). The majority of LAs (91.3%) showed an increase in diagnoses from Q3 2021 to Q4 2022.

## Discussion

National surveillance data show that there has been an increase in diagnoses of gonorrhoea in England from the cessation of social restrictions in summer 2021 through to the end of 2022. This increase was seen across people of all age groups, genders and sexual orientations; particularly among young people aged 15 to 24 and people who identify as heterosexual. This general increase in gonorrhoea diagnoses corresponds with reports from other countries in Europe, which have also seen increases in gonorrhoea diagnoses in 2022 amongst young and/or heterosexual people [4-8].

Whilst the number of tests has rebounded since the removal of COVID-19 restrictions, they have not returned to 2019 levels, especially with respect to the groups seeing the largest rises. Therefore, the increase in diagnoses cannot be explained by increased testing and is likely a result of a true increase in circulation of gonorrhoea within the population. Antimicrobial resistance of gonorrhoea to the first line treatment Ceftriaxone is closely monitored in England and cases remain infrequent, ruling out treatment failures as a contributary factor to the rise in gonorrhoea diagnoses [9, 10].

The increase is observed to be widespread and whilst most prominent in gonorrhoea, other bacterial STIs have also increased [1]. It remains to be seen whether this will be a short-lived increase resulting from the removal of restrictions alone, a resumption of the pre-COVID-19 pandemic steady rise, or another trend. Continued close surveillance and a better understanding of the factors leading to the rise will help to focus efforts to control gonorrhoea transmission.

## Data Availability

Data presented in the manuscript are available upon reasonable request to the authors

